# Deep learning algorithms for automatic segmentation of acute cerebral infarcts on diffusion-weighted images: Effects of training data sample size, transfer learning, and data features

**DOI:** 10.1101/2023.07.02.23292150

**Authors:** Yoon-Gon Noh, Wi-Sun Ryu, Dawid Schellingerhout, Jonghyeok Park, Jinyong Chung, Sang-Wuk Jeong, Dong-Seok Gwak, Beom Joon Kim, Joon-Tae Kim, Keun-Sik Hong, Kyung Bok Lee, Tai Hwan Park, Sang-Soon Park, Jong-Moo Park, Kyusik Kang, Yong-Jin Cho, Hong-Kyun Park, Byung-Chul Lee, Kyung-Ho Yu, Mi Sun Oh, Soo Joo Lee, Jae Guk Kim, Jae-Kwan Cha, Dae-Hyun Kim, Jun Lee, Man Seok Park, Dongmin Kim, Oh Young Bang, Eung Yeop Kim, Chul-Ho Sohn, Hosung Kim, Hee-Joon Bae, Dong-Eog Kim

## Abstract

**Background:** Deep learning-based artificial intelligence techniques have been developed for automatic segmentation of diffusion-weighted magnetic resonance imaging (DWI) lesions, but currently mostly using single-site training data with modest sample sizes.

**Objective:** To explore the effects of 1) various sample sizes of multi-site vs. single-site training data, 2) domain adaptation, the utilization of target domain data to overcome the domain shift problem, where a model that performs well in the source domain proceeds to perform poorly in the target domain, and 3) data sources and features on the performance and generalizability of deep learning algorithms for the segmentation of infarct on DW images.

**Methods:** In this nationwide multicenter study, 10,820 DWI datasets from 10 hospitals (Internal dataset) were used for the training-and-validation (Training-and-validation dataset with six progressively larger subsamples: n=217, 433, 866, 1,732, 4,330, and 8,661 sets, yielding six algorithms) and internal test (Internal test dataset: 2,159 sets without overlapping sample) of 3D U-net algorithms for automatic DWI lesion segmentation. In addition, 476 DW images from one of the 10 hospitals (Single-site dataset) were used for training-and-validation (n=382) and internal test (n=94) of another algorithm. Then, 2,777 DW images from a different hospital (External dataset) and two ancillary test datasets (I, n=50 from three different hospitals; II, n=250 from Ischemic Stroke Lesion Segmentation Challenge 2022) were used for external validation of the seven algorithms, testing each algorithm performance vs. manual segmentation gold standard using DICE scores as a figure of merit. Additional tests of the six algorithms were performed after stratification by infarct volume, infarct location, and stroke onset-to-imaging time. Domain Adaptation was performed to fine-tune the algorithms with subsamples (50, 100, 200, 500, and 1000) of the 2,777 External dataset, and its effect was tested using a) 1,777 DW images (from the External dataset, without overlapping sample) and b) 2,159 DW images from the Internal test dataset.

**Results:** Mean age of the 8,661 patients in the Training-and-validation dataset was 67.9 years (standard deviation 12.9), and 58.9% (n = 4,431) were male. As the subsample size of the multi-site dataset was increased from 217 to 1,732, algorithm performance increased sharply, with DSC scores rising from 0.58 to 0.65. When the sample size was further increased to 4,330 and 8,661, DSC increased only slightly (to 0.68 and 0.70, respectively). Similar results were seen in external tests. Although a deep learning algorithm that was developed using the Single-site dataset achieved DSC of 0.70 (standard deviation 0.23) in internal test, it showed substantially lower performance in the three external tests, with DSC values of 0.50, 0.51, and 0.33, respectively (all *p* < 0.001). Stratification of the Internal test dataset and the External dataset into small (< 1.7 ml; n = 994 and 1,046, respectively), medium (1.7-14.0 ml; n = 587 and 904, respectively), and large (> 14.0; n = 446 and 825, respectively) infarct size groups, showed the best performance (DSCs up to ∼0.8) in the large infarct group, lower (up to ∼0.7) in the medium infarct group, and the lowest (up to ∼0.6) in the small infarct group. Deep learning algorithms performed relatively poorly on brainstem infarcts or hyperacute (< 3h) infarcts. Domain adaptation, the use of a small subsample of external data to re-train the algorithm, was successful at improving algorithm performance. The algorithm trained with the 217 DW images from the Internal dataset and fine-tuned with an additional 50 DW images from the External dataset, had equivalent performance to the algorithm trained using a four-fold higher number (n=866) of DW images using the Internal dataset only (without domain adaptation).

**Conclusion:** This study using the largest DWI data to date demonstrates that: a) multi-site data with ∼1,000 DW images are required for developing a reliable infarct segmentation algorithm, b) domain adaptation could contribute to generalizability of the algorithm, and c) further investigation is required to improve the performance for segmentation of small or brainstem infarcts or hyperacute infarcts.

## Introduction

Diffusion weighted imaging (DWI) has been a critical imaging technique for the diagnosis and treatment of acute ischemic stroke because it is highly sensitive in detecting acute cerebral infarcts.^1^ DWI lesion volume^2^ and pattern^3^ can predict post-stroke functional outcomes and future cerebrovascular events. Moreover, DWI can guide acute recanalization therapy^4, 5^ by triaging patients based on their infarct volumes.

There is a real clinical need for automated segmentation of DW images. Since human segmentation of the infarct core demands time-consuming clinical expertise, multiple deep learning-based artificial intelligence techniques have been developed for automatic segmentation of DWI lesions.^6–9^ However, such techniques are critically dependent on the quantity and quality of the training-and-validation data (training data) used to build the algorithms, and most studies to date have utilized single-site training data with only modest sample sizes (Supplementary Table 1). Only a few studies have externally tested their deep learning algorithms, reporting-as expected that dice similarity coefficients (DSCs) were much higher for internal data than for external data.^10, 11^

Large-scale, multi-site training data are needed to avoid the two well-known machine learning failures: a) the failure of generalization problem that prevents a deep learning model from learning patterns that generalize to unseen data, and b) the domain shift problem where a model that performs well in the source domain proceeds to perform poorly in the target domain.^12^ However, collecting extensive imaging data from multiple centers is challenging. Labeling and annotating data are very labor-intensive processes that require thorough knowledge of neuroimaging. Specifically, regarding deep learning algorithms for DWI lesion segmentation, the training data sample size that minimizes the generalization problem and domain shift problem is not known yet.

To overcome the domain shift problem, domain adaptation, which fine-tunes the pre-trained model using source domain data by adjusting its parameters using additional training data from the target domain, has been successfully applied in various fields.^13^ However, studies exploring the effect of domain adaptation on the performance of deep learning algorithms for DWI infarct segmentation have not been reported yet. Clearly, the sample sizes of both initial training data and of the effects of target domain data both would be important variables to consider in such a study.

In this nationwide multi-center study (Figure 1), 10,820 patients’ DW images (collected consecutively from 10 university hospitals) were used to develop deep learning-based infarct segmentation algorithms. These algorithms were tested using three external datasets (n = 2,777, 50, and 250). We examined effects of 1) various sample sizes of multi-site vs. single-site training data, 2) domain adaptation, and 3) data sources and features on algorithm performance.

**Figure 1.**
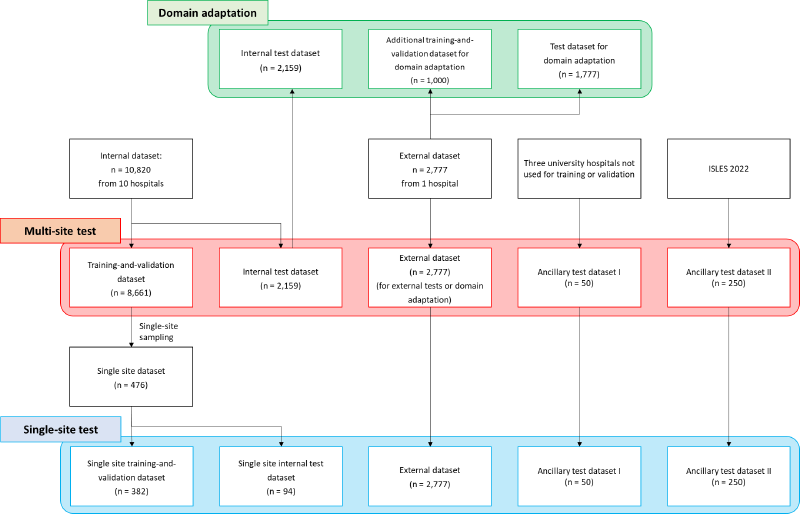
Study flow chart.

## Methods

### Training cohort

#### Multi-site data

This study included brain DW images from the Korean nationwide image-based stroke database project.^14–16^ From May 2011 to February 2014, we consecutively enrolled 12,013 patients with ischemic stroke or transient ischemic attack who were admitted to 10 stroke centers within 7 days of symptom onset. We excluded the following patients: contraindication to MRI (n = 258), poor quality or unavailability of DW images (n = 904), and MRI registration error (n = 31), leaving 10,820 patients for ‘Internal dataset’ (Figure 1). This Internal dataset was further split 80/20 into a ‘Training-and-validation dataset’ (n=8,661) and ‘Internal test dataset’ (n = 2,159).

#### Single-site data

To investigate segmentation performance of a deep learning model that was trained using data from a single site, we chose one of the 10 hospitals to prepare ‘Single-site dataset’ (Figure 1) with 476 DW images, which is comparable to the amounts of training data in previous studies.^17, 18^

### External test cohorts

Three datasets (Figure 1) were used for external validation of deep learning algorithms. First, a consecutive series of 2,777 DW images (‘External dataset’) were collected from patients who were admitted with acute ischemic stroke or transient ischemic from a university hospital during the same period as the training cohort. Second, ‘Ancillary test dataset I’ was prepared using DW images of 50 patients with ischemic stroke due to atrial fibrillation from three different university hospitals between 2011 and 2014.^19^ Third, ‘Ancillary test dataset II’ (n = 250) were Ischemic Stroke Lesion Segmentation Challenge (ISLES) 2022 data.^20^

Institutional Review Boards of all participating centers approved this study. All patients or their legally authorized representatives provided written informed consent for study participation.

### DW Images and ischemic lesion segmentation

Brain MR images for Training-and-validation dataset and internal test dataset were obtained using 1.5 Tesla (n = 6,360) or 3.0 Tesla (n = 2,882) MRI systems; data were missing in 1,259 and 319 patients for Training-and-validation dataset and internal test dataset, respectively (Supplementary Table 2). DWI protocols were: b-values of 0 and 1,000 s/mm^2^, TR of 2,400−9,000 ms, TE of 50 − 99 ms, voxel size of 1 × 1 × 3−5 mm3, interslice gap of 0−2 mm, and thickness of 3−7 mm. For the External dataset, the majority of DW images were obtained using a 1.5 Tesla MRI system (n = 2,724, 98.5%; data were missing in 12 patients). Ischemic lesions on DW images in the Training-and-validation dataset, Internal test dataset, and External dataset were segmented by experienced researchers using an in-house software Image_QNA under the close guidance by an experienced vascular neurologist, as previously described.^14, 15^ During the semi-automatic segmentation, inter-rater reliability was monitored as previously described.^21^ For the Ancillary test dataset I, an experienced neurologist manually outlined ischemic lesions. In the Ancillary test dataset II, a hybrid human-algorithm annotation scheme was applied for lesion segmentation.^20^

### Image preprocessing

To train the infarct segmentation model, brain DW images were preprocessed by (1) skull stripping using Gaussian blur and Otsu’s threshold,^22^ (2) N4 correction using the SimpleITK library, and (3) image signal image normalization as described below.

#### Skull stripping

Brain parenchyma has relatively high signal intensities in the DWI compared with skull, cerebrospinal fluid, and noisy areas. To focus on the brain parenchyma, Otsu thresholding was used to generate a parenchymal brain mask from the Gaussian blur-processed image. The brain mask was then superimposed on the original image to remove non-parenchymal structures outside the mask.

#### N4 correction

Signal intensity values of MR images are frequently non-uniform because of a bias field effect. DW images from various participating centers had different levels / distributions of signal non-uniformity. To reduce the inter-site difference, bias field correction was performed before model training, which was done using Python version of the N4 correction algorithm in the SimpleITK library. However, because the algorithm was computationally expensive, the maximum number of corrections was set to be 10 to limit the execution time for each case.

#### Image normalization

DWI signal distribution varies depending on imaging conditions such as MRI equipment vendor, magnetic field strength, echo time, and repetition time. When the noise area is removed, the peak point of the signal intensity histogram is primarily occupied by gray and white matter, with lesion and artifact areas exhibiting a higher signal, resulting in a bimodal distribution. As a normalization process to make signal intensities of each skull-stripped DW images distribute within a constant range, all the voxels in each slice was multiplied by a specific coefficient: a number found to shift the peak value in the signal intensity histogram to 150, when the peak value was divided by the number. In order to suppress abnormally high signals, which are typically noticed as artifacts in DWI,^23^ the multiplied values were clipped not to exceed 255.

### Model Development

We employed modified version of 3D U-Net^24^ for model training. While the model retained its U-shaped architecture, the number of CNN layers and the filters for these layers were modified. In addition, each convolution unit (Conv3D-BatchNormalization-ReLU) was replaced with pre-activation unit (BatchNormalization-ReLU-Conv3D), which was first utilized to increase ResNet performance^25^ and was expected to be able to boost the performance of our models.

To develop multi-site deep learning models and compare segmentation performances as training data increased, the Training-and-validation dataset was subsampled by a factor of 2.5/5/10/20/50/100% (217, 433, 866, 1,732, 4,330, and 8,661 DW images, respectively; Supplementary Fig 1), with an 8:2 training-to-validation set ratio. To develop a single-site deep learning model, a total of 476 patients’ DW images were divided into 382 (for training and validation) and 94 (for internal testing). During deep learning, random augmentation was performed in real-time to increase the diversity of training datasets and to prevent overfitting: a slice-wise affine transformation, MRI (bias field) artifact simulation, an axis flip, and a gamma/contrast change. The implementation code was developed using TorchIO, a medical imaging library written in Python.^26^ Further information is available in Supplementary Material.

In addition to the aforementioned 3D U-Net, we employed vision transformer (Swin UNETR),^27^ another well-known medical image segmentation network, for deep learning (Supplementary Material).

### Model Evaluation

After training models, segmentation performance was evaluated using the Internal test dataset, External dataset, and Ancillary test datasets I and II. As a primary evaluation metric, Dice similarity coefficient (DSC) was calculated as follows:

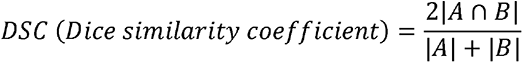

A: manual segmentation (gold standard), B: image segmentation by a deep learning algorithm

Additionally, voxel-wise sensitivity and precision were calculated by quantifying the number of missed lesion voxels or incorrectly predicted positive voxels, as follows:

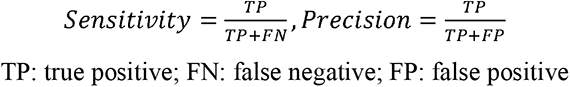

We also assessed the performance of infarct segmentation depending on the differences in:

1. - infarct volume, which was categorized as small (< 1.7 mL), medium (1.7 mL – 14 mL), and large (> 14 mL)^11^
2. - imaging acquisition time after symptom onset defined as last-known-well (< 3 hours, 3-24 hours, and > 24 hours)
3. - infarct location (cortex, corona radiata, basal ganglia and internal capsule, thalamus, midbrain, pons, medulla, cerebellum, and multiple)
4. - MRI vendor
5. - the presence vs. absence of chronic infarct, which was defined as a) 3–15 mm ischemic lesions outside the basal ganglia, brainstem, thalamus, internal capsule, or cerebral white matter or b) ischemic lesions larger than 15 mm in any areas on fluid-attenuated inversion recovery images^28^
6. - and the volume of underlying white matter hyperintensity (WMH), which was quantified as previously described^21^ and classified into deciles

### Domain adaptation

To investigate whether domain adaptation using target domain data as additional training data after initial deep learning affects segmentation performance of a fine-tuned algorithm, we randomly divided the External dataset to 1,000 images (Additional training-and-validation dataset for domain adaptation) and 1,777 images (Test dataset for domain adaptation) (Figure 1 and Supplementary Fig 2). The Additional training-and-validation dataset for domain adaptation and the Test dataset for domain adaptation were split so that there was no overlapping sample between them. The sample size for the fine tuning (i.e., additional training and validation) of the initially trained model was increased from 50 to 100, 200, 500, and 1,000 to assess the effect of domain adaptation-related data sample size on segmentation performance. The subsampled data were split at a ratio of 8:2 for training and validation. We calculated the mean and standard deviation of the DSC for both the Internal test dataset and the Test dataset for domain adaptation. Moreover, to evaluate whether the sample size of initial training dataset affects the model’s performance after domain adaptation, initial deep learning was performed with 2.5 / 5 / 10 / 20 / 50 / 100% of the Training-and-validation dataset and then fine-tuned with the Additional training-and-validation dataset for domain adaptation (sample size of 50, 100, or 200).

### Statistical analysis

To compare baseline characteristics of the Training-and-validation dataset, Internal test dataset, and External dataset, we used ANOVA, the Kruskal-Wallis test, and the chi-square test as appropriate for continuous variables and categorical variables, respectively. We used Bland-Altman plots and a linear regression analysis to compare ground truth infarct volume and segmented infarct volume by the model. To test whether DSC increased as the training sample size increased and to compare infarct volumes segmented by deep learning and manual segmentation, we used a linear regression analysis. Performance difference between models was tested using paired t-test. P-values less than 0.05 was considered statistically significant.

## Results

### Baseline characteristics of study population

Mean age of patients was 67.9 (standard deviation 12.9) years in the Training-and-validation dataset (n = 8,661). Males accounted for 58.9% (n = 4,431) (Table 1). Median NIHSS score was 4 (interquartile range 2–9) and median infarct volume was 1.95 mL. Mean age of patients was 68.2 ± 12.7 years in the Internal test dataset and 68.2 ± 12.4 years in the External dataset. Males accounted for 60.4% and 58.0% in the Internal test dataset and the External dataset, respectively. Compared with the Training-and-validation dataset and the Internal test dataset, External dataset was characterized by more cardioembolic strokes, shorter time intervals from last-known-well to imaging acquisition, and larger infarct volumes. Moreover, MR vendors, magnetic strengths, and imaging parameters were different among the Training-and-validation dataset, Internal test dataset, and External dataset (Table 1 and Supplementary Table 2). Estimated background noise and estimated signal-to-noise ratios in the Internal dataset varied widely among the 10 participating hospitals (Supplementary Fig 3).

**Table 1.**
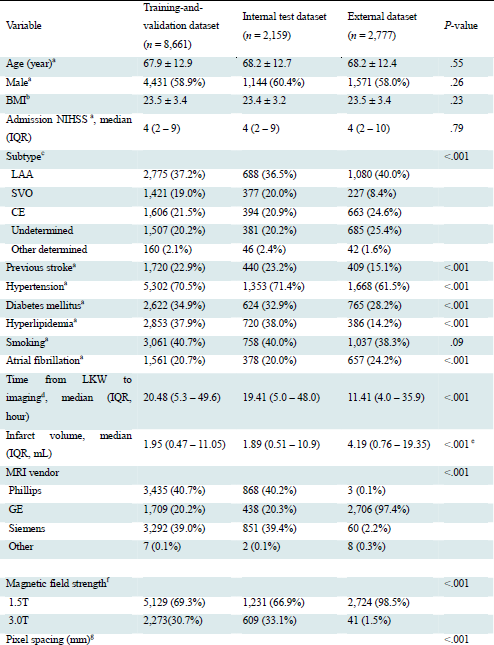
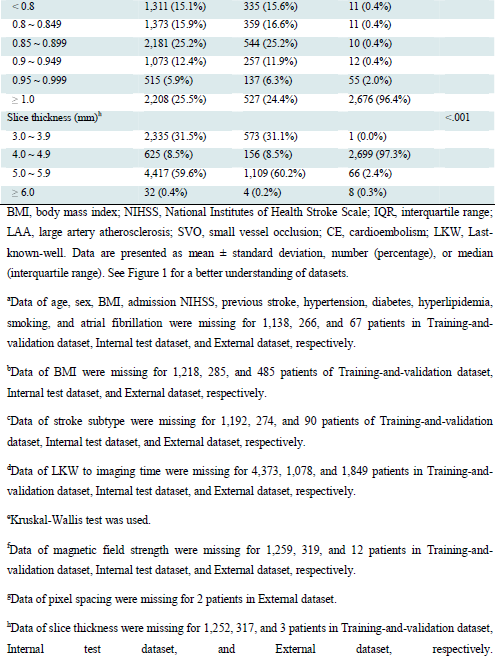
Baseline demographic and imaging characteristics of subjects, whose diffusion weighted magnetic resonances images were used for the Training-and-validation dataset, Internal test dataset, or External dataset.

### Performance of a deep learning algorithm trained using data of a single-center

To develop a single-center deep learning model, we used 382 DW images from a single hospital for model training and validation. Mean age was 68.8 ± 13.2 years in the Single site training-and-validation dataset. Males accounted for 60.8%. Median infarct volume was 1.70 (0.53–11.25) mL (Supplementary Table 3). For the Single site internal test dataset, the 3D U-net model achieved DSC of 0.70 ± 0.23 with a per-pixel sensitivity of 0.69 and a precision of 0.78 (Supplementary Table 4). However, the single-center model showed substantially lower performance for the tests using the External dataset and the Ancillary test datasets I and II, with DSC values of 0.50, 0.51, and 0.33, respectively (all *p* < 0.001).

### Effect of training data sample size on the performance of deep learning algorithm to segment acute infarcts on DW images

As the sample size of the Training-and-validation dataset increased from 217 to 433 and 866, DSC of the 3D U-net algorithm increased sharply from 0.58 to 0.61 and 0.65 for the Internal test dataset (Figure 2A). When the sample size was further increased to 1,732, DSC seemed to increase less steeply, nearly reaching a plateau (0.67). When the sample size was further increased to 4,330 and 8,661, DSC only slightly increased to 0.68 and 0.70, respectively. Similar results were seen in the tests using the External dataset (see also Supplementary Fig 4 for the Ancillary test datasets I and II). When the sample size was 433 or greater, DSC values in External dataset were significantly higher than those in Internal test dataset. In both Internal test dataset and External dataset, infarct volumes that were segmented and quantified by the 3D-Unet algorithm (trained with 8,661 DWI data) showed strong correlations with ground truth infarct volumes (both r^2^ = 0.96, *p* < 0.001; Supplementary Fig 5), although the deep learning algorithm tended to underestimate infarct volumes. Voxel-wise detection sensitivity showed a pattern that was comparable to that shown for DSCs except for fewer differences between Internal test dataset and External dataset (Figure 2B). Contrary to the exponential increase in DSC and sensitivity, precision values in both Internal test dataset and External dataset changed only slightly when training data sample size increased (Figure 2C).

**Figure 2.**
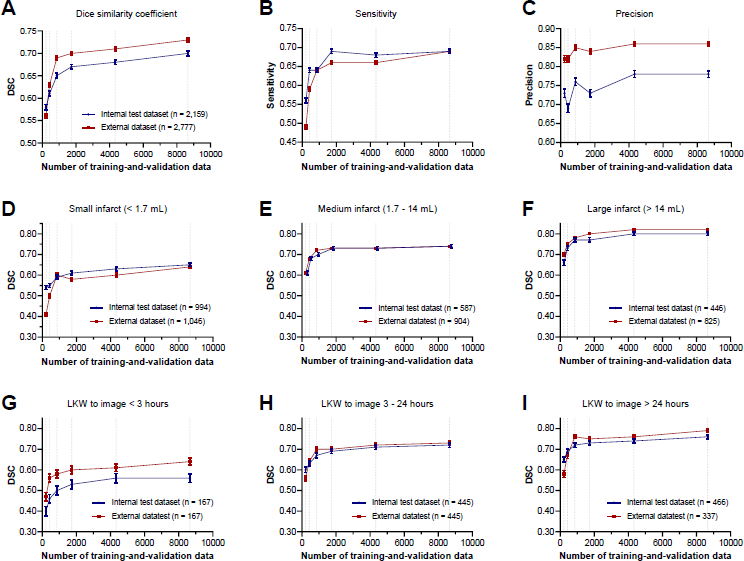
Lesion segmentation performance of deep learning algorithm as training data increase with stratification by infarct volumes and onset-to-imaging time. (A) Dice similarity coefficient (DSC) in all patients. (B) Pixel-level sensitivity in all patients. (C) Pixel-level precision in all patients. (D-F) DSC stratified by infarct volume (< 1.7, 1.7 – 14, and ≥ 14 mL). (G-H) DSC stratified by time from last-known-well to image time. Dot and bar indicate mean and standard error, respectively. Data of time from onset to imaging were missing for 565 and 1,849 patients in Internal test dataset and External dataset, respectively. Gray dot lines indicate data points of 217, 433, 866, 1,732, 4,330, and 8,661. Sensitivity and precision were calculated voxel-wise. Compared with DSC in the model trained with 217 patients, all DSCs in the model trained with 433, 866, 1,732, 4,330, and 8,661 were significantly higher. See Figure 1 for a better understanding of datasets. LKW = last-known-well.

### Effect of training data sample size on performance of deep learning algorithm to segment acute infarcts on DW images according to infarct volume, infarct location, presence of chronic ischemic lesions, onset-to-imaging time, and MRI vendors

When the Internal test dataset and the External dataset were divided into small (< 1.7 ml, n = 994 and 1,046), medium (1.7 – 14.0 ml, n = 587 and 904), and large (> 14.0, n = 446 and 825) infarct groups, DSCs for the internal and external testing were the highest (up to ∼0.8) in the large infarct group, lower (up to ∼0.7) in the medium infarct group, and the lowest (up to ∼0.6) in the small infarct group (Figure 2D-F). This finding is consistent with generally higher performances of our deep learning models in the tests using the External dataset as opposed to the Internal test dataset, given that the mean infarct volume in the former was about two times bigger than in the latter.

With regards to lesion locations (Figure 3), DSCs were generally higher for supratentorial lesions (∼0.65 or higher) than for infratentorial lesions (∼0.6 or lower), except for cerebellar lesions (in the tests using the Internal test dataset and the External dataset) and thalamus (in the test using the External dataset) with DSCs being about 0.7.

**Figure 3.**
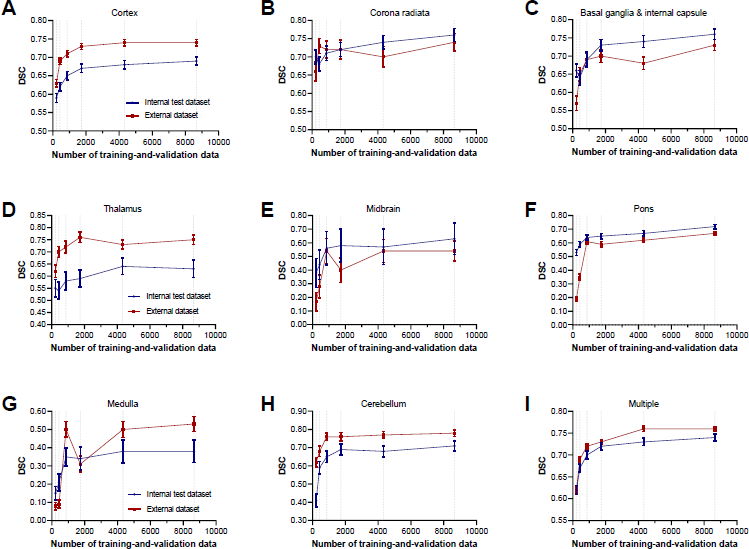
Lesion segmentation performance in the Internal test dataset and the External dataset with stratification by lesion location. (A) Cortex. (B) Corona radiata. (C) Basal ganglia & internal capsule. (D) Thalamus. (E) Midbrain. (F) Pons. (G) Medulla. (H) Cerebellum. (I) Multiple. Dot and bar indicate mean and standard error, respectively. Gray dot lines indicate data points of 217, 433, 866, 1,732, 4,330, and 8,661. Sensitivity and precision were calculated voxel-wise. Note that Y-axis ranges varied in each figure. Compared with supratentorial lesions (A-C), infratentorial lesion except for cerebellum had lower dice similarity coefficient (DSC). See Figure 1 for a better understanding of datasets.

**Figure 4.**
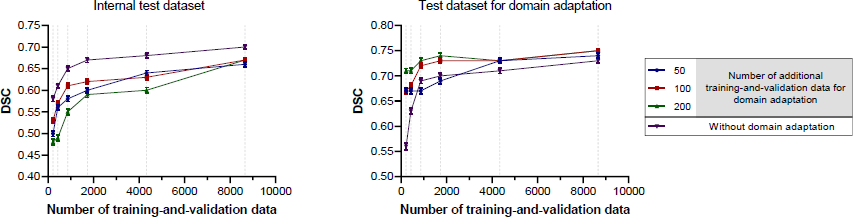
Lesion segmentation performance before and after domain adaptation using the Training-and-validation dataset for domain adaptation. (A) Dice similarity coefficient (DSC) in Internal test dataset. (B) DSC in Test dataset for domain adaptation. Data are presented as mean and stranded error. Gray dot lines indicate data points of 217, 433, 866, 1,732, 4,330, and 8,661. See Figure 1 for a better understanding of datasets.

When data were divided based on the presence of chronic ischemic lesions and WMH volumes, similar model performances were observed across groups (Supplementary Fig 6 and 7).

When data were divided based on the time from last-known-well to imaging, DSCs were the highest (up to ∼0.75) in the > 24-hour group, slightly lower (up to ∼0.7) in the 3–24-hour group, and the lowest (up to ∼0.55 and ∼0.65) in the < 3-hour group (Figure 2G-I). With respect to MRI vendors, the deep learning model showed better performances for Phillips or GE images than for Siemens images in both tests using the Internal test dataset and the External dataset (Supplementary Table 5).

In tests of the 3D-Unet model trained with 8,661 DW images, DSCs for the Internal test dataset varied, ranging from 0.45 to 0.78, depending on the participating center and training data sample size, especially the latter (Supplementary Table 6). When we employed the Swin UNETR for training with the same data, the performance of the deep learning model was generally lower than that using the 3D-Unet (Supplementary Table 7).

### Improvement of the external test performance of deep learning algorithms via domain adaptation

Domain adaptation using subsamples of the External dataset (target domain) enhanced the model performance in terms of DSC, voxel-wise sensitivity, and precision of lesion segmentation in testing with Test dataset for domain adaptation (Table 2 and Figure 3). When the sample size of the Training-and-validation dataset (source domain) was 217, retraining with 50 cases that were randomly selected from the Additional training-and-validation dataset for domain adaptation significantly increased DSC from 0.56 to 0.67 (p < 0.001; Figure 3) in testing with the Test dataset for domain adaptation. When the domain adaptation was performed with 200 cases, DSC was higher (0.71) than that for the 50 cases (p < 0.001). A similar pattern of domain adaptation-mediated performance improvement of the deep learning algorithm was observed when the sample size of the Additional training-and-validation dataset was 433. However, when the sample size was higher than 433 (i.e., 866 or higher), there was only slight improvement of infarct segmentation after domain adaptation. Thus, in terms of the effectiveness of deep learning algorithms, the training data sample size of 866 without domain adaptation was practically similar to that of 50 with subsequent domain adaptation. It is notable that domain adaptation with subsamples of target domain worsened the model performance in internal testing (i.e., testing with the source domain data). This deterioration could be partly restored by increasing the sample size of the source domain data for initial deep learning to as high as 8,661.

**Table 2.** Lesion segmentation performance after domain adaptation using the Training- and-validation dataset for domain adaptation.

| Metric | Before domain adaptation | After domain adaptation |  |  |  |  |
| --- | --- | --- | --- | --- | --- | --- |
|  |  | 50 cases | 100 cases | 200 cases | 500 cases | 1000 cases |
| Dice similarity coefficient |  |  |  |  |  |  |
| Internal test dataset ($n = 2,159$) | 0.70 (0.25) | 0.66 (0.26) | 0.67 (0.24) | 0.67 (0.24) | 0.68 (0.24) | 0.67 (0.24) |
| $P$-value<sup>a</sup> | Reference | < .001 | < .001 | < .001 | .007 | < .001 |
| Test dataset for domain adaptation ($n = 1,777$) | 0.73 (0.21) | 0.74 (0.21) | 0.75 (0.19) | 0.75 (0.19) | 0.75 (0.19) | 0.76 (0.19) |
| $P$-value<sup>a</sup> | Reference | .15 | .002 | .002 | .002 | .002 |
| Sensitivity<sup>b</sup> |  |  |  |  |  |  |
| Internal test dataset ($n = 2,159$) | 0.69 (0.27) | 0.69 (0.30) | 0.73 (0.26) | 0.71 (0.26) | 0.72 (0.25) | 0.72 (0.26) |
| $P$-value<sup>a</sup> | Reference | >.99 | < .001 | .001 | < .001 | < .001 |
| Test dataset for domain adaptation ($n = 1,777$) | 0.69 (0.23) | 0.73 (0.24) | 0.74 (0.21) | 0.73 (0.21) | 0.75 (0.21) | 0.75 (0.21) |
| $P$-value<sup>a</sup> | Reference | < .001 | < .001 | < .001 | < .001 | < .001 |
| Precision<sup>b</sup> |  |  |  |  |  |  |
| Internal test dataset ($n = 2,159$) | 0.78 (0.21) | 0.72 (0.22) | 0.68 (0.24) | 0.69 (0.25) | 0.70 (0.24) | 0.69 (0.25) |
| $P$-value<sup>a</sup> | Reference | < .001 | < .001 | < .001 | < .001 | < .001 |
| Test dataset for domain adaptation ($n = 1,777$) | 0.86 (0.16) | 0.82 (0.16) | 0.80 (0.20) | 0.82 (0.19) | 0.82 (0.19) | 0.82 (0.19) |
| $P$-value<sup>a</sup> | Reference | < .001 | < .001 | < .001 | < .001 | < .001 |

Data are presented as mean (standard deviation). See Figure 1 for a better understanding of datasets.
<sup>a</sup> $P$ -value for difference compared to the value of before domain adaptation.
<sup>b</sup>Sensitivity and precision were computed voxel-wise.

## Discussion

In the study using the largest DWI data to date, we demonstrated that the performance of 3D U-Net model for the automatic segmentation of acute infarcts improved steeply with training data volume as sample size was increased from 217 to 866 but reached a plateau as the training data was further increased to 1,732. When single-center training data was used, the performance of the deep learning algorithm degraded dramatically in external testing. Furthermore, we found that domain adaptation utilizing small amount of data from the target domain improved segmentation accuracy significantly, making the sample size of 866 without domain adaptation equivalent to that of 217 with domain adaptation.

The performance of the deep learning-based DWI lesion segmentation algorithm that was trained on the single-center dataset (n = 382) was much inferior in all three external tests than in the internal test (DSCs of 0.50, 0.51, and 0.33 vs. 0.70, respectively). To develop a more robust algorithm that generalizes well and performs better on an unseen data, there is a need for multi-site training data, which better reflects the heterogeneity of the ischemic stroke phenotype as well as the diversity of MR equipment and protocols in real-world clinical use. However, it is challenging to obtain, label, and annotate a high volume of multi-center data. Our findings suggests that multi-site data with a sample size of about 866 ∼ 1732 might be cost-effective in developing a reasonable deep learning algorithm for DWI lesion segmentation.

To enhance the deep learning model’s capacity to generalize to new cases, data augmentation can be used to artificially increase the amount and diversity of training data by generating modified copies of a dataset using existing data. However, this method carries the biases of the existing data, such as noise and resolution-related ones, without increasing the variety of infarct locations and patterns.^29^

Utilizing a small data from the target domain could be used to resolve the domain shift issue, where the model performs poorly on the target data acquired from a different source or domain (and unseen during training) due to differences in the data distributions.^12, 30, 31^ Our study showed that on the External dataset, the algorithm that was trained with 217 DW images and was followed by domain adaptation with 50 additional DW images from target domain performed comparably to the model trained with 866 DW images without subsequent domain adaptation. As a trade-off due to diversion of the deep learning model on the target domain, domain adaptation may result in worse performance in the source domain. However, resilience was observed with little impact on the model’s performance in the source domain when employing a large multi-site data for training. The post-domain-adaptation (n = 200) DSC drop for the source domain internal test data was 0.10 and 0.03, respectively, in the models that were pretrained with 866 DW images and 8,661 DW images.

Dice coefficients for DWI lesion segmentation were low when infarcts were small or MRI was performed early (within 3 hours of symptom onset). Given that the External dataset (for external testing) had approximately 2-fold bigger infarct volumes than the Internal test dataset, this finding is in line with higher DSCs for the former (vs. the latter) dataset. In addition, training on multi-site data may have led to the robustness to external testing. Deep learning algorithms performed poorly on brainstem infarcts, probably due to small number of cases even in the large training data (n=8,661) and a relatively complex anatomical structures and variations of the posterior fossa near the brainstem.^32^ A strategy for enhancing the segmentation performance for brainstem infarcts should be developed in future research.

This study has strengths, such as the large sample size of multi-site training data and extensive external test. There are also limitations. First, using apparent diffusion coefficient images for training may have enhanced the segmentation performance. Second, the performance of the algorithm may have been improved by using clinical data for training, as physicians do in clinical practice. Third, caution should be taken when extrapolating our findings from Korean stroke patients to other ethnic groups, although previous research found no ethnic differences in the pattern of ischemic infarct on DW images.^33^

In conclusion, our study demonstrates that domain adaptation or big (n=∼1000) multi-site DWI data are required for a reliable infarct segmentation algorithm with generalizability. In addition, future research should focus on improving the relatively low segmentation performance for small or brainstem infarcts or hyperacute infarcts, which has not been previously described.

## Data Availability

All data produced in the present study are available upon reasonable request to the authors.

## Acknowledgements

The authors appreciate the contributions of all members of the Comprehensive Registry Collaboration for Stroke in Korea to this study. This study was supported by the National Priority Research Center Program Grant (NRF-2021R1A6A1A03038865), the Basic Science Research Program Grant (NRF-2020R1A2C3008295), the Multiministry Grant for Medical Device Development (KMDF_PR_20200901_0098), and the Bioimaging Data Curation Center Program Grant (2022M3H9A2083956) of the National Research Foundation, funded by the Korean government.

## Tables and Figure legends for

Deep learning algorithms for automatic segmentation of acute cerebral infarcts on diffusion-weighted images: Effects of training data sample size, transfer learning, and data features (Noh et al.)

