## Supplementary Material for "Deep learning algorithms for automatic segmentation of acute cerebral infarcts on diffusion-weighted images: Effects of training data sample size, transfer learning, and data features"

| **Supplementary Method** | 2 |
| --- | --- |
| **Supplemental Figures** |  |
| Supplementary Figure 1. Experimental design testing effects of training sample size on deep learning models | 4 |
| Supplementary Figure 2. Experimental design for testing the effects of domain adaptation on performance of deep learning model | 5 |
| Supplementary Figure 3. Background noise and signal to noise ratio in the Internal dataset with stratification by participating site | 6 |
| Supplementary Figure 4. Lesion segmentation performance of 3D U-net algorithm for the Ancillary test dataset I and the Ancillary test dataset II | 7 |
| Supplementary Figure 5. Volume correlation plot between ground truth and estimated infarct volume in the Internal test dataset and the External dataset | 8 |
| Supplementary Figure 6. Lesion segmentation performance stratified by the presence of chronic ischemic lesions | 9 |
| Supplementary Figure 7. Lesion segmentation performance stratified by white matter hyperintensity volume load | 10 |
| **Supplemental Tables** |  |
| Supplementary Table 1. Summary of prior studies on deep learning algorithms to segment ischemic lesions on diffusion-weighted images | 11 |
| Supplementary Table 2. Characteristics of MRI vendors and protocols used for the Training-and-validation dataset, the Internal test dataset, and the External dataset | 13 |
| Supplementary Table 3. Baseline characteristics of the patients for the Single site training-and-validation dataset | 15 |
| Supplementary Table 4. Lesion segmentation performance of 3D U-net algorithm trained using the Single site dataset | 16 |
| Supplementary Table 5. Lesion segmentation performance depending on MRI vendors | 17 |
| Supplementary Table 6. Segmentation performance in the Internal test dataset with stratification by participating centers | 18 |
| Supplementary Table 7. Comparison of model performance between U-Net and Vision Transformer | 19 |
| **Supplemental References** | 20 |

**Supplementary Method**

**Model Development**

*3D U-Net.* The model consisted of an encoder/decoder layer that could perform max pooling four times, with feature size set to be 12, 24, 48, 96, and 192 in each step (Supplementary Fig. 1). The convolutional layer's kernel size was set to be 3 x 3 x 3. Conv3d model weights were initialized with He normal^1^ and ConvTranspose3d model weights and Xavier uniform.^2^ For model training, Adam optimizer^3^ with cyclical learning rate was used. The batch size was set to 2, and the experimental results showed that the optimal learning rate interval was 1e-5 to 1e-4. In addition, to address class imbalance caused by a small lesion size (vs. total brain volume), focal Tversky loss^4^ was used. The gamma of the loss function was set to 4/3, while alpha and beta were experimentally set to 0.6 and 0.4, respectively.

*Swin UNETR.* MONAI,^5^ a pytorch-based open source library, was used to train the model. The model's Swin Transformer set the number of transformer layers for each attention step to two and numbers of heads to two, four, eight, and sixteen. The model's weights were initialized using He normalization and focal Tversky loss function. For training the model, an Adam optimizer with a cyclical learning rate between 3e-4 and 3e-3 was used.

**Data Augmentation**

Each image has been augmented with a linear combination of the following methods.

*Slice-wise similarity transformation.* Due to the voxel anisotropy of the input image, the transformation was conducted on each slice with a randomly defined affine matrix to prevent artifacts resulting from inter-slice operations. For 2D transformation, the matrix is calculated as follows:

$$X=a*x-b*y+c$$

$$Y=b*x+a*y+d$$

$$a=s*\cos\theta$$

$$b=s*\sin\theta,$$

where $(x,y)$ and $(X,Y)$ are coordinates in the original and transformed image spaces respectively, $s$ is the scaling factor, $\theta$ is the degree of rotation, and $(c,d)$ is the translation vector for the original image. The implementation code was written using scikit-image, an imaging toolkit for Python.

*MRI bias field artifact simulation.* The bias field artifact has been modeled as a linear combination of polynomial basis functions for each axis,^6-8^ as described by the authors of TorchIO. The final simulated image is calculated as follows:

$$S=I*exp\left( BF \right)$$

where $I$ and $S$ are the original and simulated image, and $BF$ is a randomly generated bias field. To train our model, the order of the polynomial function for each axis was set to 2.

*Axis flip.* After similarity transformation and bias field simulation, a random axis flip was conducted. One of the x, y, and z axes of each image was reversed with 75% probability, and the probability of flipping each axis was the same.

*Gamma/contrast change.* Adjustment of gamma was performed by nonlinearly transforming the signal value of each voxel with an image-specific coefficient. Here, the coefficient was calculated by taking the natural exponent of a random real number between -0.3 and 0.3. The following formula is used to augment these data:

$$V_{adjusted}={V_{original}}^{\gamma},$$

where $V$ is the voxel signal and $\gamma$ is a value between $e^{-0.3}$ and $e^{0.3}$.

**Technical Details**

Python 3.7.9/3.8.13, pytorch 1.12.0, torchvision 0.13.0, pandas 1.2.4, numpy 1.19.5/1.22.3, scipy 1.4.1/1.6.3, scikit-image 0.15.0/0.18.1, SimpleITK 2.1.1, and pydicom 2.1.2 were used for all procedures, including preprocessing and model development. Intel Xeon Silver 4314 @2.40GHz, 640GB RAM, and NVIDIA Quadro RTX A6000 48GB GDDR6 were used to train models.

**Supplementary Figures**

**Supplementary Figure 1. Experimental design testing effects of training sample size on deep learning models**

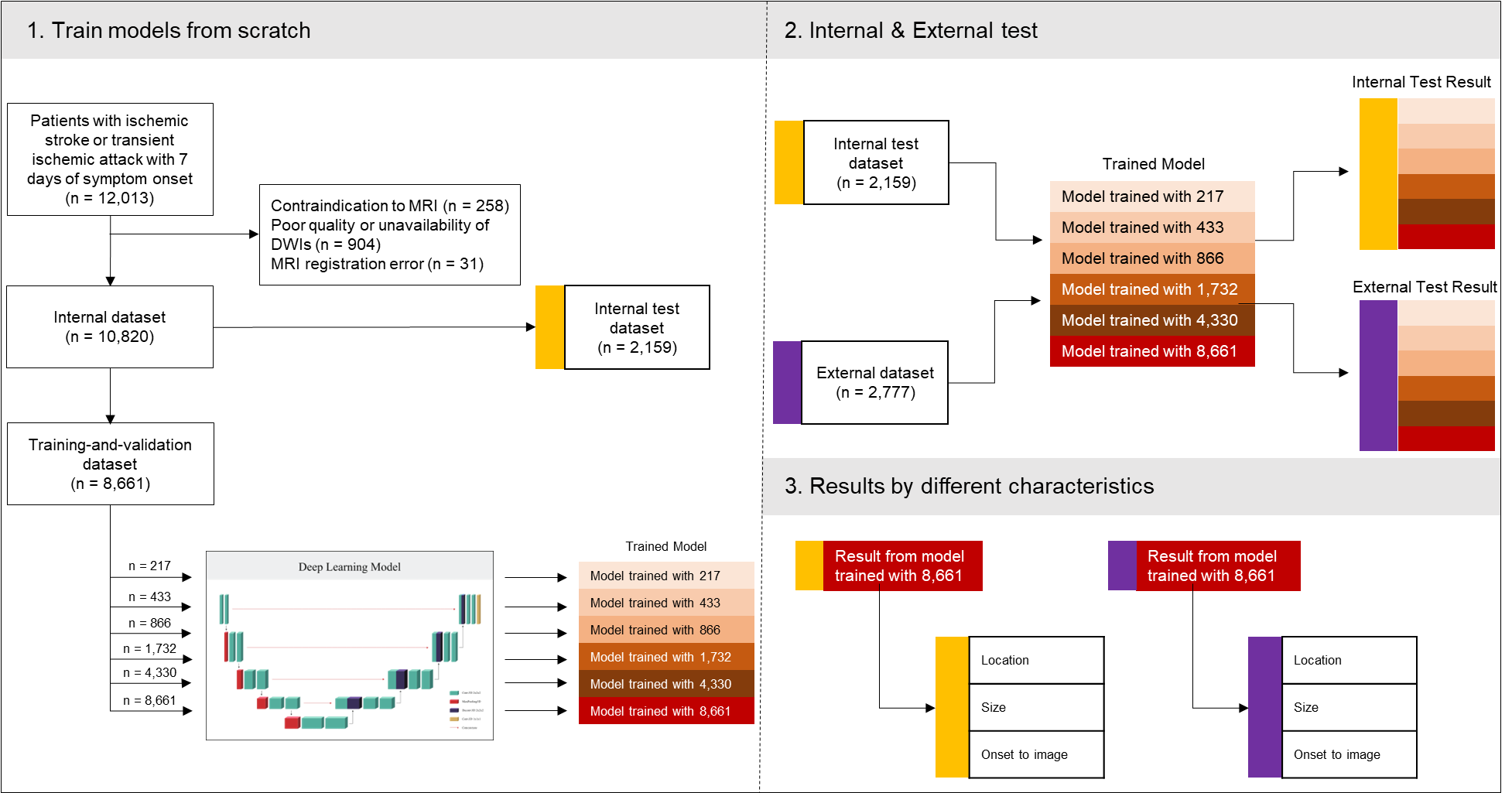

Red boxes represent baseline models trained with various sample sizes. White boxes tagged with yellow and violet markers in “2. Internal & External Test” represent the Internal test dataset and the External dataset. Red boxes tagged with yellow and violet markers indicate internal and external test results, respectively. TIA=transient ischemic stroke; MRI=magnetic resonance imaging; DWI=diffusion weighted imaging. See Figure 1 (main text) for a better understanding of datasets.

**Supplementary Figure 2. Experimental design for testing the effects of domain adaptation on performance of deep learning model**

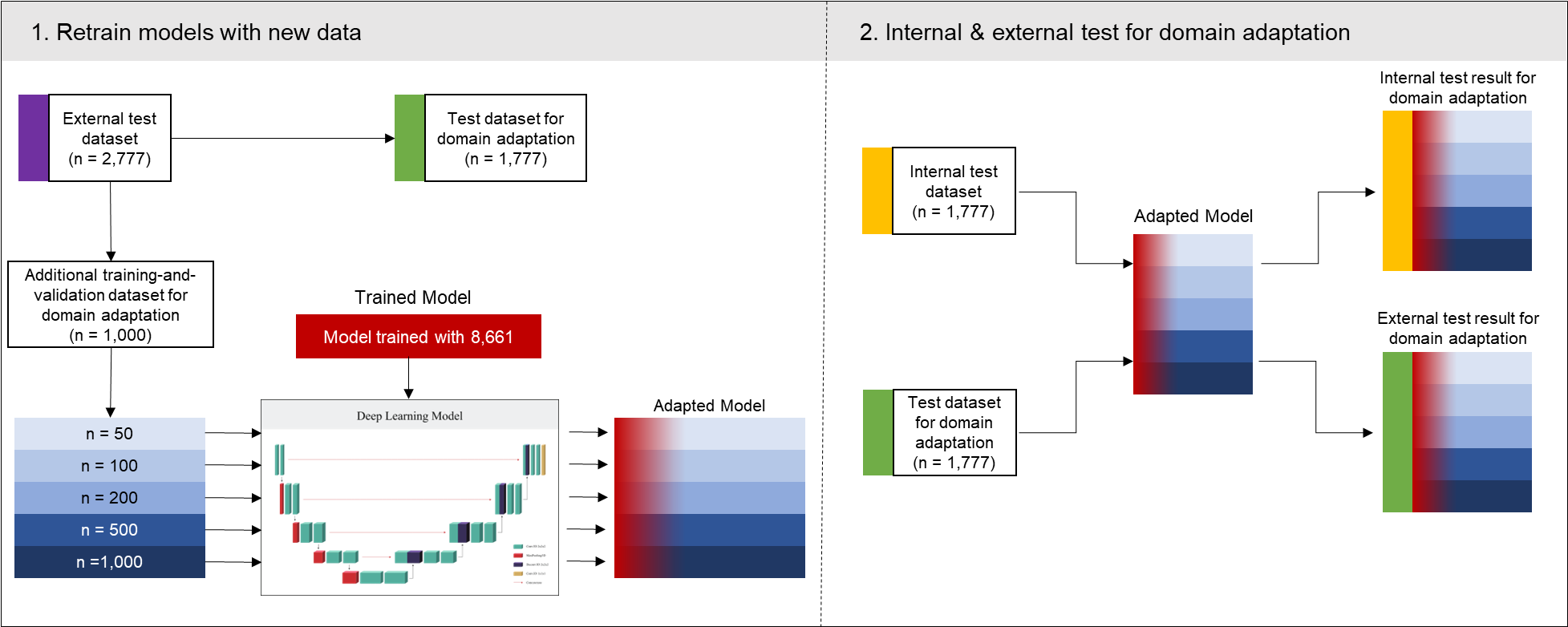

Flowchart of the process for testing the impact of sample size for domain adaptation on lesion segmentation model. Blue boxes indicate the Additional training-and-validation dataset for domain adaptation with various sample sizes. Red tinted boxes indicate baseline models trained on the source domain data. See Figure 1 (main text) for a better understanding of datasets.

**Supplementary Figure 3. Background noise and signal to noise ratio in the Internal dataset with stratification by participating site**

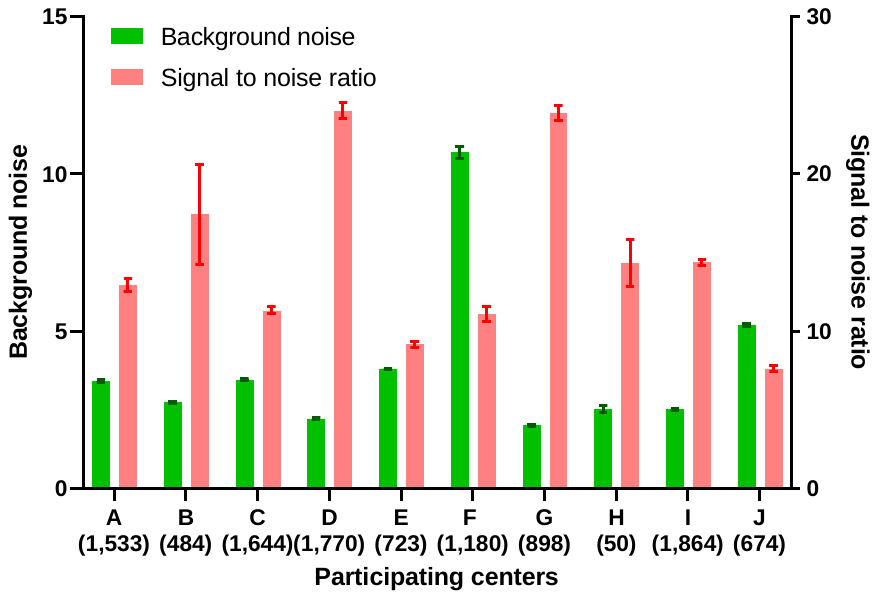

Data are depicted as mean and standard deviation. Capital letters below X-axis indicate each participating center. Numbers below capital letters indicate number of patients in each center. Background noise and signal to noise ratio were significantly different among sites (*p* < 0.001 by ANOVA). See Figure 1 (main text) for a better understanding of datasets.

**Supplementary Figure 4. Lesion segmentation performance of 3D U-net algorithm for the Ancillary test dataset I and the Ancillary test dataset II**

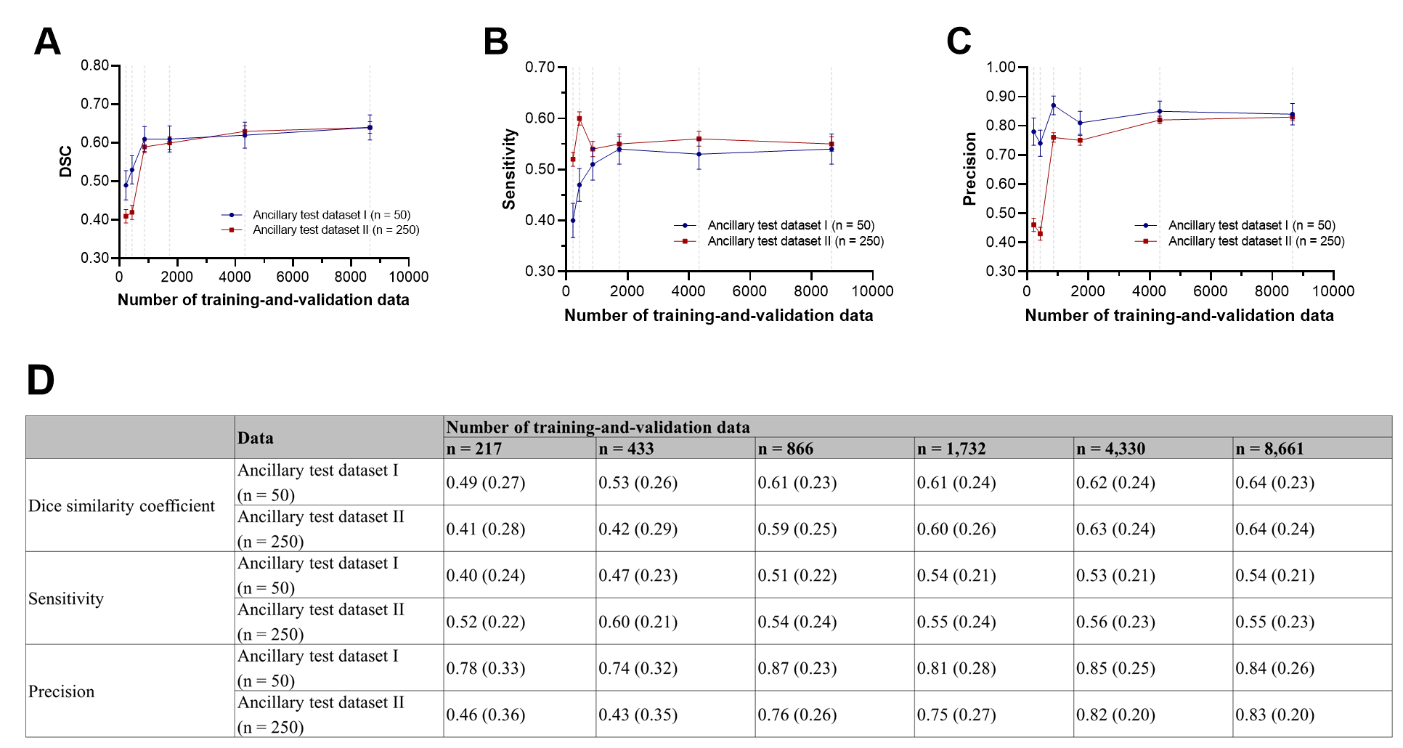

(A) Dice similarity coefficient (DSC). (B) Pixel-level sensitivity. (C) Pixel-level precision. (D) DSC, sensitivity, and specificity (SD) at each number of training-and-validate data. Data are presented as mean and stranded error. Gray dot lines indicate data points of 217, 433, 866, 1,732, 4,330, and 8,661. Sensitivity and precision were calculated voxel-wise. See Figure 1 (main text) for a better understanding of datasets.

**Supplementary Figure 5. Volume correlation plot between ground truth and estimated infarct volume in the Internal test dataset and the External dataset**

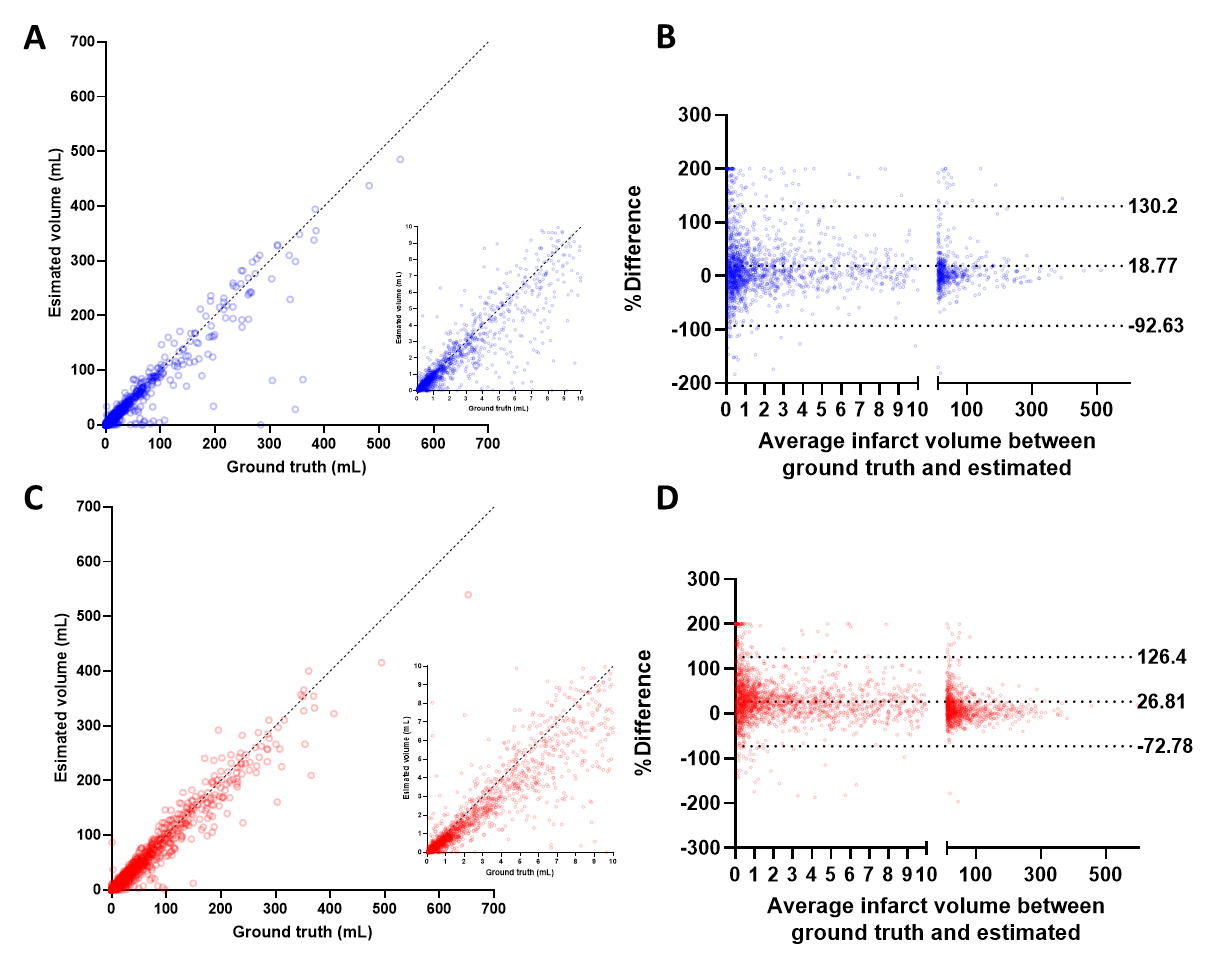

Scatter (A) and Bland-Altman (B) plots of ground truth infarct volume and deep learning estimated infarct volume in internal test data. Scatter (C) and Bland-Altman (D) plots of ground truth infarct volume and deep learning estimated infarct volume in the External dataset. In scatter plots, dotted lines indicate an indemnity line (x = y). In Bland and Altman plots dot lines indicate mean percent difference and its 95% confidence intervals. See Figure 1 (main text) for a better understanding of datasets.

**Supplementary Figure 6. Lesion segmentation performance stratified by the presence of chronic ischemic lesions**

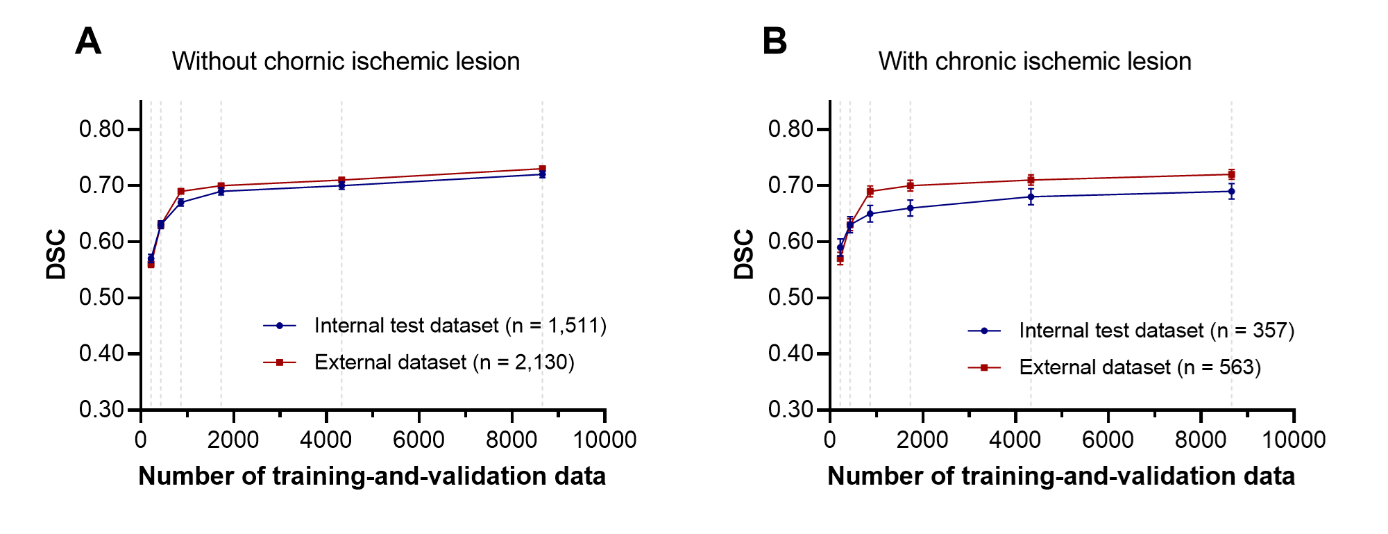

Chronic ischemic lesions on fluid-attenuated inversion recovery images were defined as a) 3–15 mm ischemic lesions outside the basal ganglia, brainstem, thalamus, internal capsule, or cerebral white matter or b) ischemic lesions larger than 15 mm in any area. Data are presented as mean and stranded error. Gray dot lines indicate data points of 217, 433, 866, 1,732, 4,330, and 8,661. See Figure 1 (main text) for a better understanding of datasets.

**Supplementary Figure 7. Lesion segmentation performance stratified by white matter hyperintensity volume load**

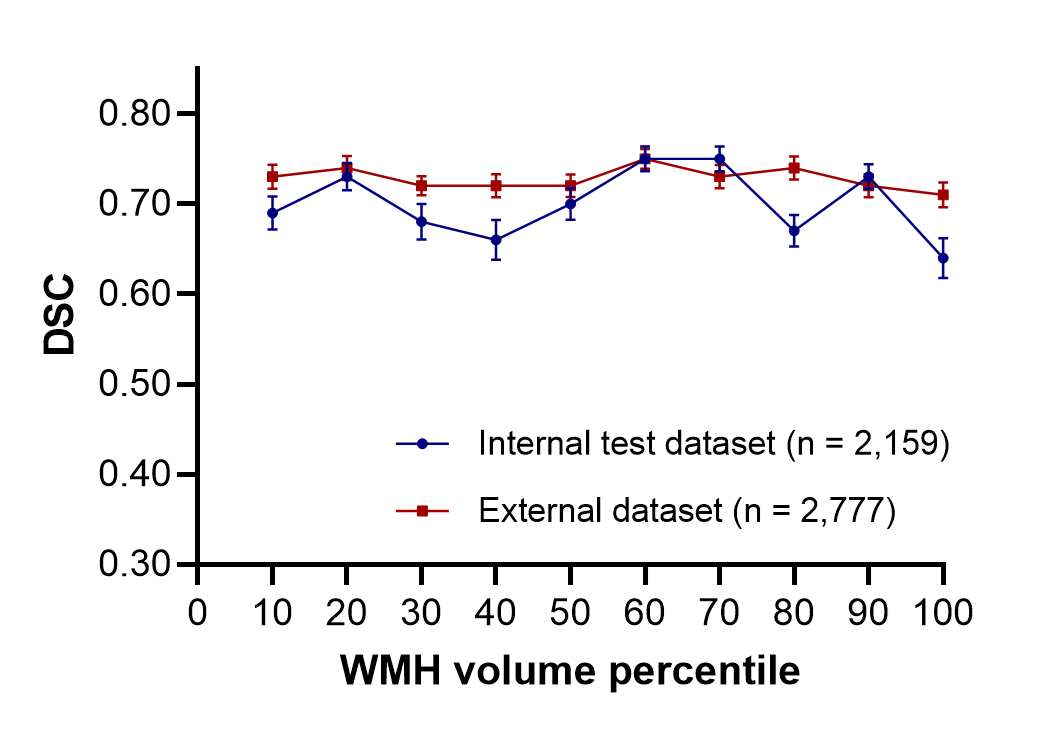

DSC = Dice similarity coefficient; WMH = white matter hyperintensity. See Figure 1 (main text) for a better understanding of datasets.

**Supplementary Tables**

**Supplementary Table 1. Summary of prior studies on deep learning algorithms to segment ischemic lesions on diffusion-weighted images**

| Ref. | No. of training & tuning data | Data source | Deep learning model | MRI vendors | External validation | Internal DSC | External DSC |
| --- | --- | --- | --- | --- | --- | --- | --- |
| [1] | 700 | Single | U-net | Siemens, GE | No | 0.85 | NA |
| [2] | 380 | Multiple | Ensemble of DeconvNets and MUSCLE Net | Siemens | No | 0.67 | NA |
| [3] | 335 | Single | Ensemble of U-net and Densenet | Siemens, GE, Philips | No | 0.86 | NA |
| [4] | 296 (test set included) | Single | U-net | Siemens, GE, Philips | No | 0.60 | NA |
| [5] | 152 | NA | 3D Densenet | Siemens, Philips | ISLES2015 | 0.79 | 0.55 |
| [6] | 1,390 | Single | DAGMNet_CH3 | Siemens, GE, Philips, other | STIR2 | 0.76 | 0.51 |
| [7] | 116 | Single | DeepMedic | GE | No | 0.82 | NA |
| [8] | 115 | Single | Res-FCN | Siemens | ISLES2015 | 0.80 | 0.65 |
| [9] | 460 | Single | DPC-Net | Siemens | No | 0.64 | NA |
| [10] | 2385 and 3157 | Multiple (6) | U-net | Siemens, GE | Cross-vendor test^a^ | 0.87 and 0.90 | 0.73 and 0.76 |

DSC = Dice similarity coefficient; ISLES = Ischemic Stroke Lesion Segmentation; STIR = Stroke Imaging Repository; NA = not available

^a^Models were independently trained using either Siemens or GE data and externally validated using other vendors’ data.

**Supplementary Table 2. Detailed characteristics of MRI vendors and protocols used for the Training-and-validation dataset, the Internal test dataset, and the External dataset**

| Variable | Training-and-validation  (*n* = 8,661) | Internal test dataset  (*n* = 2,159) | External dataset  (*n* = 2,777) | *P*-value |
| --- | --- | --- | --- | --- |
| MRI model |  |  |  | <.001 |
| Phillips – Achieva | 642 | 167 | 0 (0.0%) |  |
| Phillips – Intera | 2,063 | 512 | 0 (0.0%) |  |
| Phillips – Others | 730 | 189 | 3 (0.1%) |  |
| GE-Signa Excite | 1,263 | 303 | 4 (0.1%) |  |
| GE-Signa HDxt | 19 | 7 | 1,494 (53.8%) |  |
| GE-Discovery MR750 | 414 | 121 | 4 (0.1%) |  |
| GE-Others | 13 | 7 | 1,204 (43.4%) |  |
| Siemens-Avanto | 1,554 | 364 | 4 (0.1%) |  |
| Siemens-Sonata | 684 | 166 | 0 (0.0%) |  |
| Siemens-Skyra | 757 | 202 | 0 (0.0%) |  |
| Siemens-Others | 297 | 119 | 56 (2.0%) |  |
| Other MRI vendors | 7 | 2 | 8 (0.3%) |  |
| Magnetic field strength^a^ |  |  |  | <.001 |
| 1.5T | 5,129 | 1,231 | 2,724 (98.5%) |  |
| 3.0T | 2,273 | 609 | 41 (1.5%) |  |
| Acquisition matrix (row) |  |  |  | <.001 |
| < 192 | 1,539 | 360 | 46 (1.7%) |  |
| 192 ~ 239 | 657 | 154 | 10 (0.4%) |  |
| 240 ~ 255 | 633 | 165 | 0 (0.0%) |  |
| 256 ~ 319 | 4,860 | 1,234 | 2,710 (97.6%) |  |
| 320 ~ 511 | 607 | 155 | 11 (0.4%) |  |
| 512 | 365 | 91 | 0 (0.0%) |  |
| Pixel spacing (mm)^b^ |  |  |  | <.001 |
| < 0.8 | 1,311 | 3335 | 11 (0.4%) |  |
| 0.8 ~ 0.849 | 1,373 | 359 | 11 (0.4%) |  |
| 0.85 ~ 0.899 | 2,181 | 544 | 10 (0.4%) |  |
| 0.9 ~ 0.949 | 1,073 | 257 | 12 (0.4%) |  |
| 0.95 ~ 0.999 | 515 | 137 | 55 (2.0%) |  |
| ≥ 1.0 | 2,208 | 527 | 2,676 (96.4%) |  |
| Slice thickness (mm)^c^ |  |  |  | <.001 |
| 3.0 ~ 3.9 | 2,335 | 573 | 1 (0.0%) |  |
| 4.0 ~ 4.9 | 625 | 156 | 2,699 (97.3%) |  |
| 5.0 ~ 5.9 | 4,417 | 1,109 | 66 (2.4%) |  |
| ≥ 6.0 | 32 | 4 | 8 (0.3%) |  |
| Repetition time (ms)^d^ |  |  |  | <.001 |
| < 3000 | 613 | 142 | 2 (0.1%) |  |
| 3000 ~ 3999 | 2,465 | 600 | 10 (0.4%) |  |
| 4000 ~ 4999 | 1,618 | 403 | 6 (0.2%) |  |
| 5000 ~ 5999 | 1,065 | 278 | 43 (1.5%) |  |
| 6000 ~ 6999 | 617 | 154 | 1 (0.0%) |  |
| 7000 ~ 7999 | 545 | 134 | 10 (0.4%) |  |
| ≥ 8000 | 1,717 | 444 | 2,705 (97.4%) |  |
| Echo time (ms) |  |  |  | <.001 |
| < 50 | 66 | 11 | 0 (0.0%) |  |
| 50 ~ 59.99 | 874 | 224 | 0 (0.0%) |  |
| 60 ~ 69.99 | 1,931 | 478 | 0 (0.0%) |  |
| 70 ~ 79.99 | 2,885 | 726 | 7 (0.3%) |  |
| 80 ~ 89.99 | 994 | 248 | 2,685 (97.5%) |  |
| 90 ~ 99.99 | 1,117 | 281 | 49 (1.8%) |  |
| ≥ 100 | 756 | 175 | 14 (0.5%) |  |
| FOV (row, mm) |  |  |  | <.001 |
| < 210 | 644 | 153 | 5 (0.2%) |  |
| 210 ~ 219 | 1,816 | 464 | 11 (0.4%) |  |
| 220 ~ 229 | 2,893 | 713 | 10 (0.4%) |  |
| 230 ~ 239 | 2,238 | 562 | 54 (1.9%) |  |
| 240 ~ 249 | 984 | 238 | 16 (0.6%) |  |
| 250 ~ 259 | 57 | 14 | 56 (2.0%) |  |
| ≥ 260 | 29 | 15 | 2,623 (94.5%) |  |

MRI, magnetic resonance imaging; FOV, field of view.

^a^Data of magnetic field strength were missing for 1,259, 319, and 12 patients in the Training-and-validation dataset, Internal test dataset, and External dataset, respectively. See Figure 1 (main text) for a better understanding of datasets.

^b^Data of pixel spacing were missing for 521, 132, and 2 patients in the Training-and-validation dataset, the Internal test dataset, and the External dataset, respectively.

^c^Data of slice thickness were missing for 1,251, 317, and 3 patients in the Training-and-validation dataset, the Internal test dataset, and the External dataset, respectively.

^d^Data of repetition time were missing for 21 and 4 patients in the Training-and-validation dataset and the Internal test dataset, respectively.

**Supplementary Table 3. Baseline characteristics of the patients for the Single site training-and-validation dataset**

| Variable | Single site training-and-validation dataset (*n* = 382) |
| --- | --- |
| Age (year)^a^ | 68.8 ± 13.2 |
| Male^a^ | 225 (60.8%) |
| BMI^a^ | 23.0 ± 3.6 |
| Admission NIHSS^a^, median (IQR) | 4 (2-8) |
| Subtype^a^ |  |
| LAA | 135 (36.5%) |
| SVO | 95 (25.7%) |
| CE | 67 (18.1%) |
| Undetermined | 62 (16.8%) |
| Other determined | 11 (3.0%) |
| Previous stroke^a^ | 102 (27.6%) |
| Hypertension^a^ | 278 (75.1%) |
| Diabetes mellitus^a^ | 137 (37.0%) |
| Hyperlipidemia^a^ | 184 (49.7%) |
| Smoking^a^ | 155 (41.9%) |
| Atrial fibrillation^a^ | 64 (17.3%) |
| Time from LKW to imaging^a^, median (IQR, hour) | 19.02 (3.7-50.8) |
| Infarct volume^a^, median (IQR, mL) | 1.70 (0.53-11.25) |

BMI, body mass index; NIHSS, National Institutes of Health Stroke Scale; IQR, interquartile range; LAA, large artery atherosclerosis; SVO, small vessel occlusion; CE, cardioembolism; LKW, Last known well.

Data are presented as mean ± standard deviation, number (percentage), or median (interquartile range). See Figure 1 (main text) for a better understanding of datasets.

^a^Data of age, sex, BMI, admission NIHSS, subtype, previous stroke, hypertension, diabetes, hyperlipidemia, smoking, atrial fibrillation, LKW to imaging time, and infarct volume were missing for 12 patients.

**Supplementary Table 4. Lesion segmentation performance of 3D U-net algorithm trained using the Single site dataset**

| Metric | Single site internal test dataset  (*n* = 94) | External dataset  (*n* = 2,777) | Ancillary test dataset I  (*n* = 50) | Ancillary test dataset II  (*n* = 250) |
| --- | --- | --- | --- | --- |
| Dice similarity coefficient | 0.70 (0.23)  [0.64, 0.85] | 0.50 (0.31)  [0.21, 0.76] | 0.51 (0.26)  [0.27, 0.72] | 0.33 (0.30)  [0.05, 0.60] |
| *P* for difference | Reference | < .001 | < .001 | < .001 |
| Sensitivity | 0.69 (0.25)  [0.61, 0.87] | 0.46 (0.31)  [0.18, 0.72] | 0.43 (0.23)  [0.35, 0.58] | 0.59 (0.21)  [0.47, 0.74] |
| *P* for difference | Reference | < .001 | < .001 | < .001 |
| Precision | 0.78 (0.18)  [0.71, 0.89] | 0.68 (0.34)  [0.52, 0.94] | 0.76 (0.32)  [0.65, 0.98] | 0.34 (0.37)  [0.03, 0.71] |
| *P* for difference | Reference | .005 | .63 | < .001 |

The model was trained and validated using 382 patients from a single center. Results are presented as mean (standard deviation) and [interquartile range]. Sensitivity and precision were calculated voxel-wise. See Figure 1 (main text) for a better understanding of datasets.

**Supplementary Table 5. Lesion segmentation performance depending on MRI vendors**

|  | Number of training-and-validation data | | | | | |
| --- | --- | --- | --- | --- | --- | --- |
| MRI Vendor | *n* = 217 | *n* = 433 | *n* = 866 | *n* = 1,732 | *n* = 4,330 | *n* = 8,661 |
| Phillips |  |  |  |  |  |  |
| Internal test dataset (n = 868) | 0.62 (0.28) | 0.64 (0.27) | 0.68 (0.26) | 0.69 (0.26) | 0.71 (0.24) | 0.73 (0.24) |
| External dataset (n = 3) | N/A | N/A | N/A | N/A | N/A | N/A |
| GE |  |  |  |  |  |  |
| Internal test dataset (n = 438) | 0.55 (0.29) | 0.62 (0.26) | 0.65 (0.26) | 0.67 (0.26) | 0.68 (0.26) | 0.70 (0.24) |
| External dataset (n = 2,703) | 0.56 (0.28) | 0.63 (0.27) | 0.70 (0.23) | 0.70 (0.24) | 0.71 (0.23) | 0.73 (0.21) |
| Siemens |  |  |  |  |  |  |
| Internal test dataset (n = 851) | 0.55 (0.29) | 0.59 (0.28) | 0.62 (0.28) | 0.64 (0.26) | 0.65 (0.27) | 0.66 (0.25) |
| External dataset (n = 60) | 0.48 (0.31) | 0.50 (0.31) | 0.58 (0.31) | 0.61 (0.28) | 0.64 (0.28) | 0.64 (0.28) |

Results are presented as mean (standard deviation) of Dice similarity coefficient. See Figure 1 (main text) for a better understanding of datasets.

^a^Data are either missing or coming from other vendors for 2 and 11 patients in the Internal test dataset and External test dataset, respectively.

**Supplementary Table 6. Segmentation performance in the Internal test dataset with stratification by participating centers**

|  | Number of training-and-validation samples | | | | | |
| --- | --- | --- | --- | --- | --- | --- |
|  | *n* = 217 | *n* = 433 | *n* = 866 | *n* = 1,732 | *n* = 4,330 | *n* = 8,661 |
| Hospital 1  (n = 307) | 0.57 (0.28) | 0.62 (0.26) | 0.67 (0.25) | 0.68 (0.26) | 0.69 (0.26) | 0.72 (0.24) |
| Hospital 2  (n = 96) | 0.59 (0.31) | 0.66 (0.27) | 0.70 (0.25) | 0.68 (0.27) | 0.71 (0.25) | 0.71 (0.25) |
| Hospital 3  (n = 329) | 0.64 (0.28) | 0.68 (0.24) | 0.70 (0.24) | 0.71 (0.23) | 0.73 (0.23) | 0.74 (0.23) |
| Hospital 4  (n = 354) | 0.58 (0.29) | 0.59 (0.28) | 0.64 (0.29) | 0.66 (0.27) | 0.68 (0.26) | 0.69 (0.26) |
| Hospital 5  (n = 145) | 0.65 (0.24) | 0.72 (0.19) | 0.76 (0.15) | 0.76 (0.17) | 0.78 (0.15) | 0.78 (0.15) |
| Hospital 6  (n = 236) | 0.45 (0.31) | 0.45 (0.32) | 0.51 (0.32) | 0.57 (0.29) | 0.55 (0.32) | 0.59 (0.29) |
| Hospital 7  (n = 180) | 0.65 (0.29) | 0.68 (0.25) | 0.72 (0.26) | 0.73 (0.25) | 0.75 (0.23) | 0.77 (0.20) |
| Hospital 8  (n = 10) | 0.61 (0.29) | 0.61 (0.27) | 0.69 (0.24) | 0.68 (0.28) | 0.72 (0.27) | 0.73 (0.21) |
| Hospital 9  (n = 368) | 0.56 (0.27) | 0.60 (0.27) | 0.60 (0.28) | 0.62 (0.26) | 0.62 (0.27) | 0.64 (0.25) |
| Hospital 10  (n = 134) | 0.50 (0.29) | 0.60 (0.25) | 0.62 (0.26) | 0.66 (0.24) | 0.66 (0.24) | 0.69 (0.23) |

Results are presented as mean (standard deviation). See Figure 1 (main text) for a better understanding of datasets.

**Supplementary Table 7. Comparison of model performance between U-Net and Vision Transformer**

| Metric | U-Net | Swin UNETR |
| --- | --- | --- |
| Dice similarity coefficient |  |  |
| Internal test dataset (*n* = 2,159) | 0.70 (0.25)  [0.63, 0.86] | 0.67 (0.27)  [0.58, 0.86] |
| External dataset (*n* = 2,777) | 0.73 (0.21)  [0.68, 0.86] | 0.67 (0.27)  [0.60, 0.86] |
| Sensitivity |  |  |
| Internal test dataset (*n* = 2,159) | 0.69 (0.27)  [0.58, 0.89] | 0.67 (0.29)  [0.53, 0.89] |
| External dataset (*n* = 2,777) | 0.69 (0.23)  [0.58, 0.85] | 0.62 (0.28)  [0.48, 0.84] |
| Precision |  |  |
| Internal test dataset (*n* = 2,159) | 0.78 (0.21)  [0.71, 0.93] | 0.76 (0.24)  [0.70, 0.93] |
| External dataset (*n* = 2,777) | 0.86 (0.16)  [0.81, 0.96] | 0.87 (0.16)  [0.83, 0.87] |

Results are presented as mean (standard deviation) and [interquartile range]. Both models' training utilized 8,661 samples for training-and-validation. Sensitivity and precision were calculated voxel-wise. See Figure 1 (main text) for a better understanding of datasets.
